# Use of Envelope Following Response Normative Ranges for Diagnosing Cochlear Deafferentation

**DOI:** 10.1101/2025.10.24.25338742

**Authors:** Anne E. Heassler, Garnett P. McMillan, Sean D Kampel, Nicole K. Whittle, Haley A. Szabo, Sarah Verhulst, Brad N. Buran, Naomi F. Bramhall

## Abstract

**Purpose:** The lack of a means for diagnosing cochlear synaptopathy, a type of cochlear deafferentation, prevents clinicians from identifying patients with this auditory deficit and providing them with appropriate treatments. The envelope following response (EFR) has potential as a diagnostic indicator of deafferentation. However, it is not clear what constitutes an abnormal EFR response. The objectives of this study were to establish normative ranges for EFR magnitude in a population at low risk for cochlear synaptopathy and then compare EFRs from a population at high risk for synaptopathy to those normative ranges.

**Methods:** The low-risk sample consisted of young adults with normal audiograms, minimal reported lifetime noise exposure, and no auditory complaints. Normative ranges were generated using rectangular amplitude modulated (RAM) or sinusoidal amplitude modulated (SAM) EFR stimuli and were adjusted for sex and distortion product otoacoustic emission (DPOAE) levels. The high-risk sample consisted of military Veterans with normal audiograms who reported at least one auditory complaint (tinnitus, decreased sound tolerance, or speech-in-noise difficulty).

**Results:** The SAM EFR normative ranges for a 4 kHz carrier resulted in the biggest separation of the low-and high-risk samples, with 36% of Veterans falling below the lower bound of the normative range. There were no consistent effects of DPOAE adjustment on the normative ranges across sex and stimulus condition and computational modeling suggests that adjusting for DPOAEs may not be necessary in individuals with normal audiograms.

**Conclusion:** EFR normative ranges for the 4 kHz SAM EFR will allow for clinical identification of patients with normal audiograms who have significant degrees of cochlear deafferentation.

## Introduction

Audiologists rely on standardized audiometric tests to accurately diagnose various types of hearing loss and to determine appropriate individualized treatment plans. These tests, including tympanometry, pure tone audiometry, speech reception thresholds, and word recognition tests, have served as the standard for evaluating hearing for decades. While effective for identifying conventional hearing loss, in patients with normal audiometric test results these standard measures often fail to explain auditory complaints such as difficulty understanding speech in noisy environments, the perception of tinnitus, and reduced sound tolerance (e.g., Billings et al., 2018; Koerner et al., 2020). Notably, Tremblay et al. (2015) found that approximately 12% of individuals with normal hearing thresholds report auditory complaints, suggesting the presence of underlying auditory dysfunction in these individuals that standard audiometric tests do not detect.

One possible etiology for auditory complaints in individuals with normal hearing thresholds is cochlear synaptopathy, a type of cochlear deafferentation characterized by loss of the synaptic connections between the inner hair cells (IHCs) and the afferent auditory nerve fibers. Cochlear synaptopathy has been demonstrated in response to age and noise exposure in multiple animal models, including mouse (Buran et al., 2025; Kujawa & Liberman, 2009; Sergeyenko et al., 2013), guinea pig (Lin et al., 2011), gerbil (Schmiedt et al., 1996), rat (Niwa et al., 2016), chinchilla (Bharadwaj et al., 2022), and non-human primates (Valero et al., 2017). This suggests that synaptopathy may be a common source of auditory dysfunction in humans. Cochlear synaptopathy can result in reduced peripheral auditory input to the central auditory system without negatively impacting outer hair cells (OHCs) or audiometric thresholds (Kujawa & Liberman, 2009), making it difficult to detect clinically. Synaptopathy is expected to lead to auditory perceptual deficits including tinnitus, hyperacusis, and difficulty with speech perception in noise (Kujawa & Liberman, 2015).

Temporal bone studies indicate that age-and noise-related synaptic loss occurs in humans, independent of hair cell degeneration (Wu et al., 2019; Wu et al., 2021).

Currently, cochlear synaptopathy can only be confirmed through post-mortem temporal bone analysis. However, three auditory physiological measures that may be sensitive to cochlear deafferentation have been identified in animal models. These include auditory brainstem response (ABR) wave I amplitude (Kujawa & Liberman, 2009), the middle-ear muscle reflex (MEMR; Valero et al., 2018), and the envelope following response (EFR; Shaheen et al., 2015). It is important to note that none of these measures are specifically sensitive to cochlear synaptopathy. Instead, they are sensitive to all types of cochlear deafferentation, including loss of cochlear synapses, inner hair cells (IHCs), and spiral ganglion cells. However, human temporal bone studies suggest that synaptopathy is the most common type of cochlear deafferentation (Wu et al., 2019; Wu et al., 2021).

The EFR is an auditory evoked potential generated in response to amplitude modulated sounds. Like the ABR, it is recorded using scalp electrodes and reflects the auditory system’s ability to encode the temporal envelope of amplitude modulated sounds (Dolphin & Mountain, 1992). The EFR shows strong associations with histologically confirmed synaptopathy in animal models (Buran et al., 2025; Parthasarathy & Kujawa, 2018; Shaheen et al., 2015) and synaptopathy risk factors (age and noise exposure) in humans (Bramhall et al., 2021; Bramhall et al., 2023). Human studies also suggest that reduced EFR magnitude is associated with predicted perceptual consequences of synaptopathy, such as tinnitus and difficulty with speech-in-noise perception (Bramhall, Buran, & McMillan, 2025; Bramhall & McMillan, 2024). The modulation frequency of the EFR influences which generators contribute to the response due to phase locking limitations at higher levels of the auditory system. EFRs recorded with the modulation frequencies (f_m_) typically used in human synaptopathy studies (f_m_ =∼85-120 Hz) are thought to be dominated by generators in the auditory brainstem (Herdman et al., 2002; Kiren et al., 1994; Kuwada et al., 2002). The EFR (f_m_=1000 Hz), presumed to be generated by auditory nerve fibers, demonstrates the most sensitivity to age-and noise exposure-related synaptopathy in mice (Parthasarathy & Kujawa, 2018; Shaheen et al., 2015). Because it is difficult to get a measurable EFR (f_m_=1000 Hz) in humans, lower modulation frequencies are typically used in human studies, although recent work suggests that it is possible to measure EFR (f_m_=1024 Hz) in humans using an ear canal electrode (McHaney et al., 2024; Zink et al., 2025).

While many human studies of synaptopathy/deafferentation have used a sinusoidally amplitude modulated (SAM) EFR stimulus, computational modeling suggests that a rectangular amplitude modulated (RAM) stimulus provides enhanced neural synchrony due to the sharper rise time (Vasilkov et al., 2021). This increased synchrony leads to a stronger EFR response and may allow for greater sensitivity to cochlear deafferentation (Van Der Biest et al., 2023). Several human studies have shown a relationship between RAM EFR magnitude and speech perception performance (Bramhall & McMillan, 2024; Garrett et al., 2025; Mepani et al., 2021).

Although cochlear synaptopathy can occur in the absence of OHC damage, the risk factors for synaptopathy (age and noise exposure) are also risk factors for OHC damage. It is therefore expected that synaptopathy will often co-occur with OHC damage. Therefore, even in individuals with clinically normal hearing, it is important to consider the possible impacts of subclinical OHC dysfunction on the EFR measurement. Changes in EFR magnitude due to OHC dysfunction could confound the use of EFR magnitude as a measure of cochlear deafferentation.

While the EFR has potential as a non-invasive diagnostic indicator of cochlear deafferentation, it cannot currently be used for this purpose because there is no established normative range for EFR magnitude in humans. Normative ranges are crucial for diagnostic testing for many reasons, including establishing baseline comparisons, standardizing results, and guiding clinical decision-making. For example, pure tone threshold testing uses a normative range that was developed through a combination of early psychophysical studies of human auditory sensitivity and established clinical audiometric procedures (Carhart & Jerger, 1959; Goodman, 1965; Jerger & Jerger, 1980). Audiologists compare pure tone thresholds from individual patients to established normative ranges to determine the degree of hearing loss. Given the high expected prevalence of cochlear deafferentation, it is important for clinicians to have a diagnostic measure, with normative ranges, that they can use to identify patients with this type of auditory dysfunction.

The primary objective for this study was to develop EFR normative ranges in a population at low risk for cochlear synaptopathy based on young age (18-35 years), limited noise exposure history, and the absence of auditory complaints (tinnitus, decreased sound tolerance, or speech perception difficulties). To account for OHC dysfunction, the normative ranges were adjusted for average distortion product otoacoustic emission (DPOAE) level. The secondary objective was to compare EFR magnitudes obtained from a population at high risk for cochlear synaptopathy (military Veterans with normal audiograms who report auditory complaints) to the EFR normative ranges. It was assumed that if the magnitude of the EFR is a reliable indicator of cochlear deafferentation, many of the individuals from the high-risk sample would fall below the normative ranges.

## Methods

### Participants

Participants were recruited from the VA Portland Health Care System, local colleges and universities, and the greater Portland, OR region. All study procedures were approved by the Institutional Review Board of the VA Portland Health Care System. Informed consent was obtained from all participants prior to initiating any study activities, and participants were compensated for their time.

#### Normative low-risk sample

Data was collected from 154 young adults, aged 18 to 35 years (mean age = 24.2 years, standard deviation (SD) = 3.9 years; 44 males, 110 females), with normal hearing thresholds from 0.25 to 8 kHz (<u>≤</u>20 dB HL). These individuals reported minimal lifetime noise exposure and denied any auditory complaints, such as tinnitus, decreased sound tolerance, or difficulty with speech perception. Given their young age, limited noise exposure, and absence of auditory complaints, these participants were expected to be at low risk for cochlear synaptopathy. All participants had normal tympanograms (± 50 daPa, compliance 0.3-1.5 ml) in at least one ear. Individuals with a history of concussion were not included due to potential impacts on the EFR.

Lifetime noise exposure history was assessed in the low-risk sample using the recreational and non-military occupational sections of the Lifetime Exposure to Noise and Solvents Questionnaire (LENS-Q; Griest-Hines et al., 2021). The LENS-Q was scored as described by Griest-Hines et al. Potential participants with a LENS-Q score ≥ 5 were excluded due to high noise exposure history because a previous study in this population found a mean LENS-Q score of 4.1 with a standard deviation of 0.8 (Bramhall et al., 2021). Individuals reporting any history of military service or firearm use were excluded from this sample.

Potential participants also completed a custom questionnaire about auditory complaints. The perception of tinnitus was assessed with the question “Have you experienced ringing, roaring, or buzzing in the ears or head (tinnitus) that lasts for at least 5 minutes?” Those who reported, “yes” were excluded from the study. Self-reported difficulty hearing speech in background noise was evaluated by asking potential participants to rate their perceived difficulty hearing in various listening situations, using questions from the Hearing section of the Tinnitus and Hearing Survey (THS; Henry et al., 2015). These situations included noisy or crowded places, understanding speech on TV or in movies, understanding people with soft voices, and understanding group conversations. Individuals who reported difficulty in two or more of these situations were excluded from the sample. Potential participants also answered the following question from the Sound Tolerance section of the THS: “Over the last week, have sounds been too loud or uncomfortable for you when they seemed normal to others around you?” If they responded, “yes” they were asked to provide two examples of such sounds. If the examiner, a licensed audiologist, judged their responses as consistent with decreased sound tolerance, they were excluded from the sample.

Normative ranges for the MEMR generated from data obtained from 141 of these low-risk participants were previously reported (Bramhall, McMillan, et al., 2025).

#### High-risk comparison sample

Eighty-two military Veterans, aged 23-49 years (mean age = 38 years, standard deviation (SD) = 6.3 years; 51 males, 31 females), with normal hearing thresholds and normal tympanograms (same criteria as for the low-risk sample) who reported at least one auditory complaint made up the high-risk comparison sample. These individuals were assumed to be at high risk for synaptopathy due to their history of noise exposure during their military service and their report of auditory complaints. Veterans with a history of concussion were not included because this could impact the EFR measurements.

Veterans were asked the same questions about auditory complaints (tinnitus, difficulty hearing speech in background noise, and decreased sound tolerance) as the low-risk participants. Only Veterans who reported tinnitus, difficulty hearing in speech in at least two of the situations from the THS, and/or decreased sound tolerance were included in the study. Veteran participants also completed the Speech, Spatial, and Qualities of Hearing Scale (SSQ-12; Noble et al., 2013), a 12-item questionnaire that assesses difficulty hearing in various situations. Average scores on the SSQ-12 range from 0 to 10, with lower scores indicating greater difficulty. Veterans who reported tinnitus also completed the Tinnitus Functional Index (TFI; Meikle et al., 2012), a 25-item questionnaire that assesses the social and emotional impact of tinnitus on quality of life. TFI total score ranges from 0 to 100, with higher scores indicating greater tinnitus severity and functional impact.

MEMR data from 81 of the high-risk participants were reported previously (Bramhall et al. 2025).

## Procedures

### Audiometric Evaluation

All participants underwent a screening audiometric evaluation in both ears that included otoscopy, tympanometry, and air-and bone-conduction pure tone threshold testing. Only potential participants who met the audiometric inclusion criteria in at least one ear were invited to participate. All other study measures were obtained in a single test ear. If only one ear met the audiometric criteria, that ear was used as the test ear. Some participants had cerumen or an ear piercing in one ear that could potentially impact testing. In these cases, the other ear was used as the test ear if it also met the inclusion criteria. In all other situations where both ears qualified, the test ear was determined by rolling a die.

### Distortion Product Otoacoustic Emissions (DPOAEs)

As an indicator of outer hair cell (OHC) function, DPOAEs were recorded in the test ear using a custom system consisting of an Etymotic Research ER-10X probe microphone and EMAV software (Neely & Liu, 1993). A primary frequency sweep (DP-gram) was obtained in 1/3-octave increments from 1 to 8 kHz and 1/6-octave increments from 8 to 16 kHz, with a frequency ratio of *f_2_/f_1_* = 1.2 and primary levels set at *L_1_* = 65 dB FPL and *L_2_*= 55 dB FPL. At each frequency, data collection continued until a 48-second sample of artifact-free data was obtained, the signal-to-noise ratio (SNR) was ≥ 30 dB, or the noise floor dropped below-30 dB FPL. In-the-ear calibration was used to adjust the output voltage so that *L_1_*and *L_2_* were presented at the desired levels. The ER-10X allows forward pressure level (FPL) calibration which enhances the accuracy of stimulus level measurements, particularly in the 3-7 kHz range, where the interaction between the incident and reflected waves in the adult ear canal can create standing wave pressure nodes that can lead to measurement error when using standard SPL calibration (Konrad-Martin et al., 2016). Aside from the resulting differences in measurement error, the FPL and SPL values are equivalent.

DPOAE levels were averaged from either 3-8 kHz or 3-16 kHz to statistically adjust the EFR normative ranges for OHC function. Although the 3-16 kHz average provides a better indication of OHC function in the basal region of the cochlea, many clinicians and researchers do not have the equipment necessary to measure DPOAEs in the extended high frequencies. For this reason, the DPOAE average from 3-8 kHz was also used as an alternative. If the DPOAE level measured at any frequency was < - 20 dB SPL, the DPOAE level at that frequency was excluded from the DPOAE average.

### Envelope Following Response (EFR)

The EFR was obtained in all participants using the Intelligent Hearing Systems Smart EP system with SmartEP-CAM software (Miami, FL) in an acoustically treated sound booth with electrical power to the sound booth turned off during testing to limit contamination from the line frequency. Participants were seated in a comfortable recliner and were encouraged to sleep during testing. Electrodes were placed at Cz (active), Fpz (ground), and the mastoid of the test ear (reference). Electrode impedance was less than or equal to 5.0 kΩ in all but one participant, who had an impedance of 5.5 kΩ. Stimuli were presented through electrostatically shielded Etymotic Research ER-3A insert transducers (Grand Prairie, TX) with shielded cables. Stimuli consisted of RAM tones with carrier frequencies of 2, 4, or 8 kHz or a SAM tone with a carrier frequency of 4 kHz. All stimuli were 100% modulated (f_m_=110 Hz) and were 500 ms in duration. RAM stimuli had a 25% duty cycle and were ramped with a 2.5% tapered-cosine Tukey window. The SAM stimulus was calibrated to have a root mean square (RMS) sound pressure of 70 dB SPL averaged over multiple cycles, while the RAM stimuli were calibrated to 70 dB peak equivalent SPL (dB peSPL), measured from the positive peak to the negative peak, calculated as 20 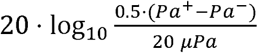 where Pa^+^ is the amplitude of the positive peak and Pa^-^ is the amplitude of the negative peak. Averaging together the positive and negative amplitudes corrects for any potential DC offset introduced by the calibration microphone. Since the RAM EFR is reported in dB peSPL, the actual level in dB RMS will be dB peSPL + 10 • log_10_ D where D is duty cycle expressed as a fraction.

For a duty cycle of 0.25, the overall level in dB SPL will be-6 dB lower than the value calculated in dB peSPL (i.e., 64 dB SPL). All stimuli were calibrated twice per year using a Brϋel & Kjær (Darmstadt, Germany) head and torso simulator (Model 4128) with artificial ear simulators (4158-C, 4159-C), and a 3160-A-042 digitizer. See **Figure 1** for waveforms and fast Fourier transforms (FFTs) of the EFR stimuli, as recorded using the head and torso simulator, artificial ear simulator, and ER-3A transducer. For the 8 kHz RAM EFR stimulus, transients were present at the onset and offset of the stimulus (**Figure 1E**), potentially an artifact resulting from the frequency response of the ER-3A transducer which rolls off for stimuli above 4 kHz. For calibration purposes, these transients were ignored. RAM stimuli were presented 1002 times and SAM stimuli were presented 3002 times because the SAM EFR has a much smaller magnitude than the RAM EFR, requiring additional averaging to get a robust EFR. It took roughly 10 minutes to collect the data from each RAM stimulus and 30 minutes to collect the data for each SAM stimulus. The acquisition sampling rate was 10,000 Hz and a bandpass filter of 100-4000 Hz at 6 dB/octave was used.

**Figure 1.**
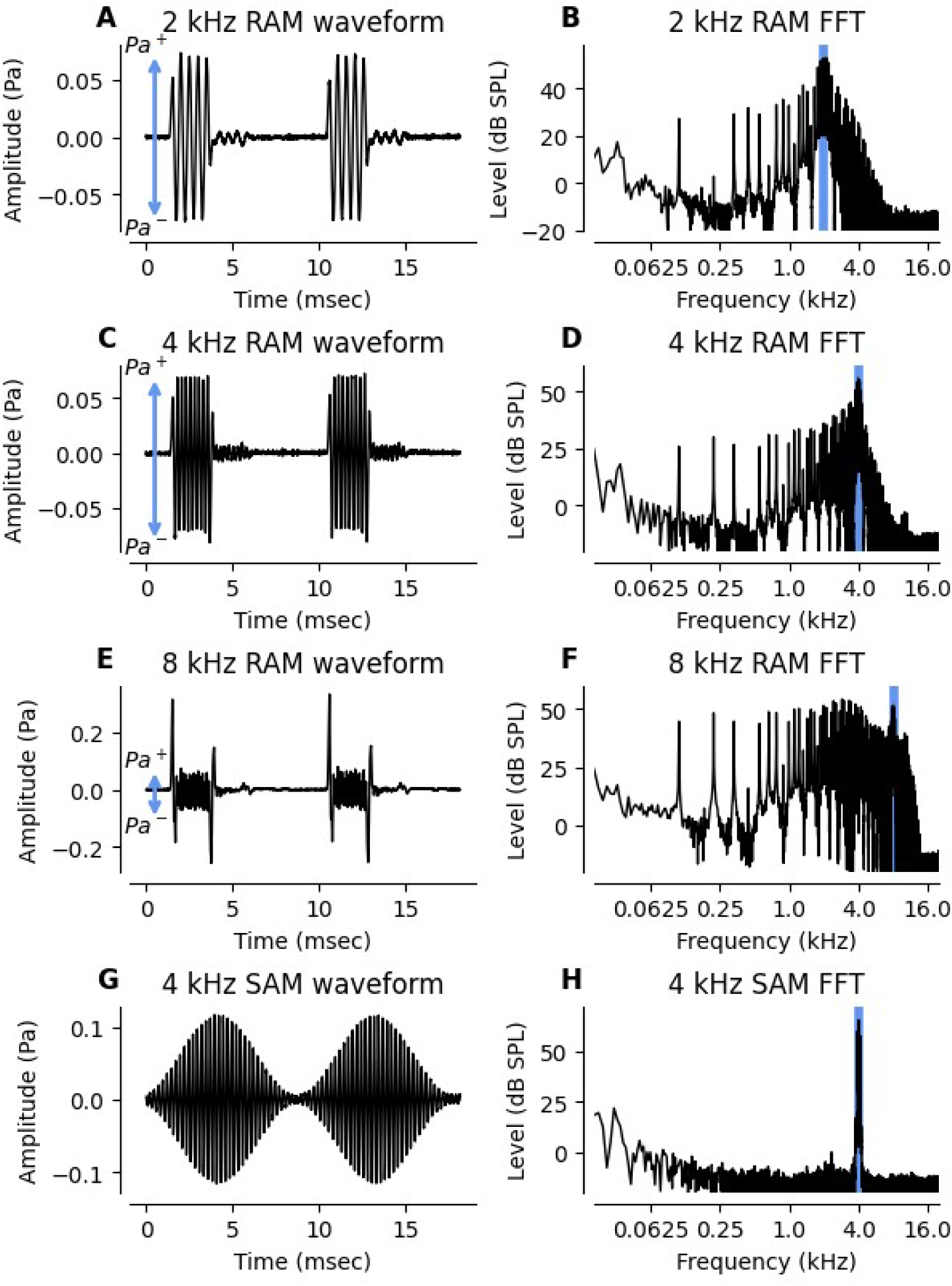
Recorded waveforms and fast Fourier transforms for RAM and SAM EFR stimuli. Stimuli were presented through an ER-3A transducer and the output was measured using a head and torso simulator with an artificial ear simulator. First column (A, C, E, G) shows the waveform of the first two cycles of the stimulus as measured by the microphone embedded in the ear simulator. For the RAM EFR (A, C, E), stimulus level was calculated by measuring the difference between the positive and negative peaks (blue arrow) and reported in dB peSPL. For the 8 kHz RAM EFR, the onset and offset transients were ignored when calculating the peak-to-peak amplitude (E). For the SAM EFR (G), the stimulus level was calculated as the root-mean-square (RMS) of the amplitude and was averaged over multiple cycles. Second column (B, D, F, H) shows the frequency spectrum of each stimulus as calculated from the full 500 msec stimulus. Blue line indicates the carrier frequency of the stimulus.

EFR magnitude was calculated similarly to the bootstrapping approach described in Zhu et al. (2013). In this approach, 400 trials were drawn with replacement, averaged, and the power spectrum was computed. Random draws were balanced across positive and inverted polarities (i.e., 200 trials from each polarity). This process was repeated 100 times to generate a distribution of the magnitude for each frequency bin. The average value of the distribution at the modulation frequency and the first four harmonics was used to estimate the raw EFR response. To estimate the noise floor, the power in the fourth to seventh discrete Fourier transform (DFT) bin on either side of the frequency of interest was averaged for a total of eight bins. EFR magnitude was calculated in dB SNR as 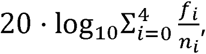 where f is the power of the i-th harmonic and n_i_ is the power of the noise floor surrounding the i-th harmonic.

### Generation of EFR Normative Ranges

Normative ranges are commonly used to identify patients with atypical features compared to a ‘healthy, normal’ population (Wright & Royston, 1999). Normative ranges represent specific percentiles of the reference population’s measurement distribution.

For example, an 80% reference interval spans the 10^th^ to 90^th^ percentiles of the measurement distribution. Measurements that fall outside of this range are considered unusual.

Conditional normative ranges are population percentiles that vary based on specific patient characteristics. Age-specific reference intervals, for example, are adjusted according to the age of the reference population and are generally estimated by collecting measurements from a large number of healthy individuals across different age groups. An alternative approach to generating conditional normative ranges involves statistical modeling of the relationship between the population percentiles and a given characteristic. This method, known as quantile regression, involves fitting regression models to estimate the 10^th^ and 90^th^ percentiles in a sample of healthy individuals with varying levels of the conditioning variable. While standard regression models focus on the mean of the measurement distribution, quantile regression allows for more flexibility when modeling the data distribution based on covariates.

In this analysis, only data from the low-risk sample contributed to the normative ranges. For each EFR condition (RAM 2 kHz, RAM 4 kHz, RAM 8 kHz, and SAM 4 kHz), the 10^th^ and 90^th^ percentiles of the EFR magnitude distribution were conditioned on participant sex and average DPOAE levels (averaged from either 3-16 kHz or 3-8 kHz). Various non-linear relationships between the percentiles and DPOAE levels were explored, including smoothing splines and quadratic models, but these approaches performed poorly in the sample and did not offer clear advantages over a linear model. The final model accounted for percentiles based on average DPOAE level, participant sex, and the interaction between DPOAE level and sex. Given that the normative ranges were developed to identify individuals likely to have high degrees of cochlear deafferentation, the lower bounds (10^th^ percentile) of the normative ranges were of particular interest because falling below the lower bound is indicative of an abnormally small EFR magnitude.

## Results

EFR normative ranges for males and females, adjusted for average DPOAE level from 3-16 kHz or 3-8 kHz, are shown in **Figure 2** and **Figure 3**, respectively (blue shaded region). The lower bounds of the normative ranges were defined by the 10^th^ percentile of the low-risk sample, and the upper bounds were defined by the 90^th^ percentile. This means that 10% of a new sample from the same low-risk population would be expected to fall below the lower bounds of the normative ranges while another 10% would be expected to surpass the upper bounds. This lower bound is the value that is of particular interest for the purpose of identifying cochlear deafferentation.

**Figure 2.**
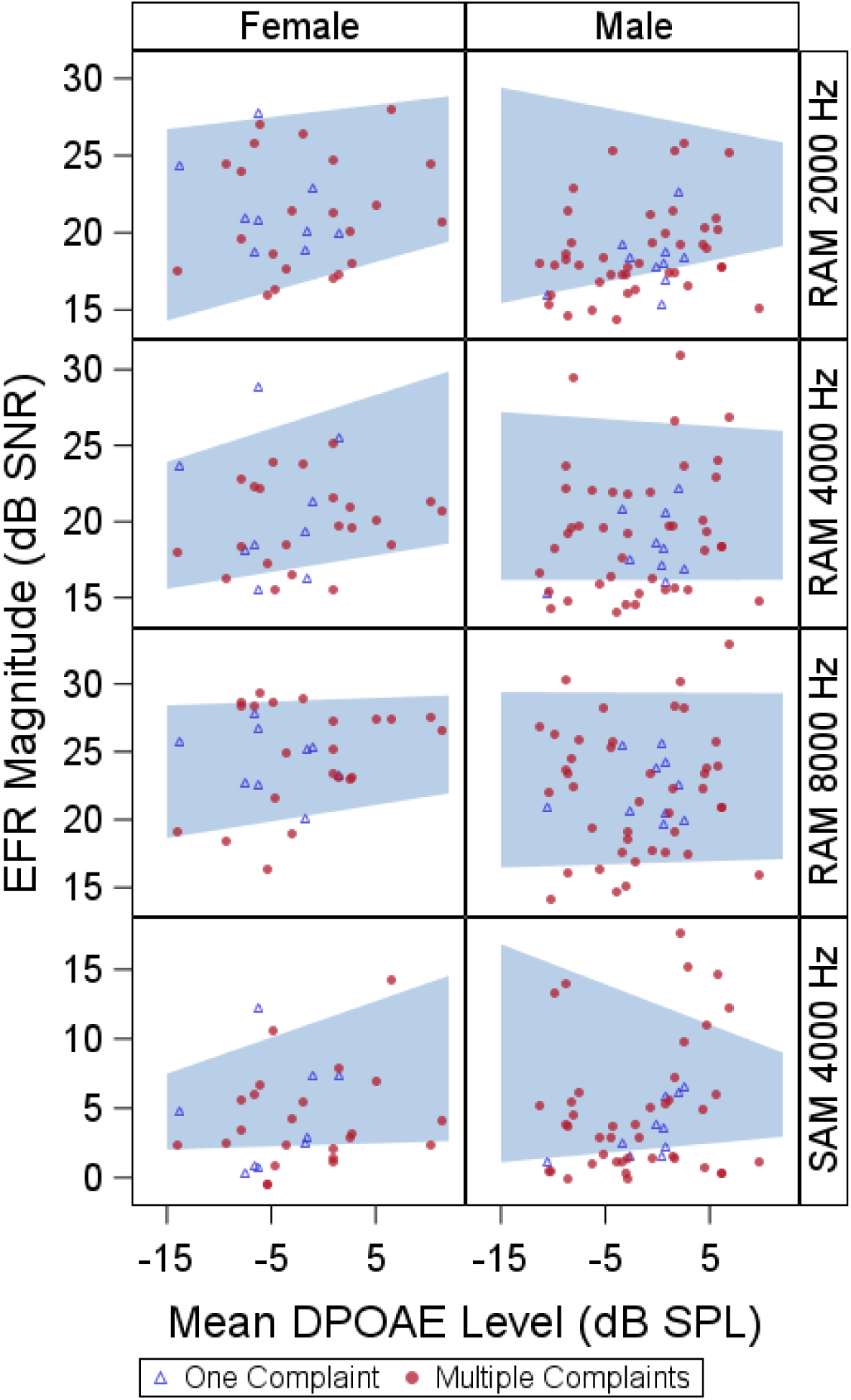
EFR normative ranges adjusted for sex and average DPOAE level from 3-16 kHz. Each panel shows the normative range for a different EFR stimulus with female normative ranges in the left column and male normative ranges in the right column. Blue shaded region indicates the DPOAE-adjusted normative range. Symbols show how EFR data from high-risk participants compare to the normative ranges (open blue triangles for individuals with one auditory complaint, filled red circles for individuals with multiple auditory complaints).

**Figure 3.**
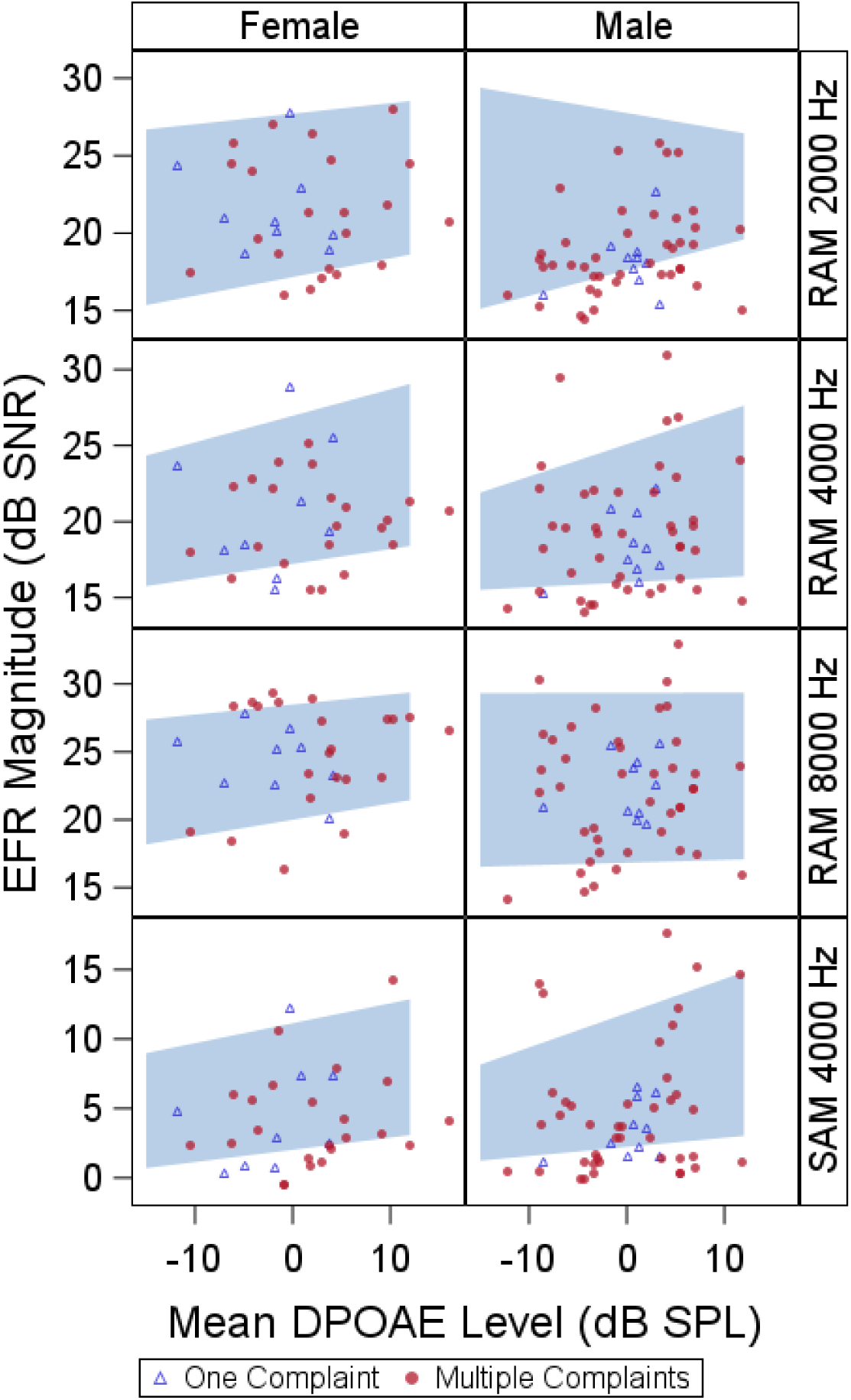
EFR normative ranges adjusted for sex and average DPOAE level from 3-8 kHz. Each panel shows the normative range for a different EFR stimulus with female normative ranges in the left column and male normative ranges in the right column. Blue shaded region indicates the DPOAE-adjusted normative range. Symbols show how EFR data from high-risk participants compare to the normative ranges (open blue triangles for individuals with one auditory complaint, filled red circles for individuals with multiple auditory complaints).

Individuals whose EFR magnitude falls below the lower bound are considered to have an abnormally small EFR magnitude. The EFR magnitudes at the upper and lower bounds of the normative ranges for different average DPOAE levels are shown in **Table 1** and **Table 2** for DPOAEs averaged from 3-16 kHz and 3-8 kHz, respectively.

**Table 1.** Lower (10^th^ percentile) and upper bounds (90^th^ percentile) for EFR normative ranges adjusted for sex and average DPOAE levels from 3-16 kHz.

| Upper bound (90<sup>th</sup> percentile) |  |  |  |  |  |  |  |  |
| --- | --- | --- | --- | --- | --- | --- | --- | --- |
|  | Male |  |  |  | Female |  |  |  |
|  | EFR Stimulus |  |  |  |  |  |  |  |
| Average DPOAE level (dB SPL) | RAM 2 kHz | RAM 4 kHz | RAM 8 kHz | SAM 4 kHz | RAM 2 kHz | RAM 4 kHz | RAM 8 kHz | SAM 4 kHz |
| -15 | 29.42 | 27.21 | 29.39 | 16.82 | 26.71 | 23.92 | 28.42 | 7.49 |
| -10 | 28.76 | 26.98 | 29.37 | 15.38 | 27.10 | 25.03 | 28.56 | 8.80 |
| -5 | 28.10 | 26.75 | 29.36 | 13.93 | 27.50 | 26.13 | 28.70 | 10.10 |
| 0 | 27.44 | 26.52 | 29.35 | 12.48 | 27.89 | 27.23 | 28.83 | 11.41 |
| 5 | 26.78 | 26.29 | 29.33 | 11.03 | 28.29 | 28.34 | 28.97 | 12.71 |
| 10 | 26.12 | 26.06 | 29.32 | 9.59 | 28.68 | 29.44 | 29.11 | 14.02 |
| Lower bound (10<sup>th</sup> percentile) |  |  |  |  |  |  |  |  |
|  | Male |  |  |  | Female |  |  |  |
|  | EFR Stimulus |  |  |  |  |  |  |  |
| Average DPOAE level (dB SPL) | RAM 2 kHz | RAM 4 kHz | RAM 8 kHz | SAM 4 kHz | RAM 2 kHz | RAM 4 kHz | RAM 8 kHz | SAM 4 kHz |
| -15 | 15.43 | 16.14 | 16.46 | 1.10 | 14.30 | 15.57 | 18.64 | 2.02 |
| -10 | 16.11 | 16.14 | 16.58 | 1.44 | 15.25 | 16.13 | 19.25 | 2.13 |
| -5 | 16.80 | 16.15 | 16.70 | 1.78 | 16.20 | 16.68 | 19.86 | 2.25 |
| 0 | 17.49 | 16.15 | 16.81 | 2.12 | 17.14 | 17.23 | 20.47 | 2.36 |
| 5 | 18.17 | 16.15 | 16.93 | 2.46 | 18.09 | 17.78 | 21.08 | 2.47 |
| 10 | 18.86 | 16.16 | 17.04 | 2.80 | 19.04 | 18.33 | 21.69 | 2.58 |

Values are in dB SNR.

**Table 2.** Lower (10^th^ percentile) and upper bounds (90^th^ percentile) for EFR normative ranges adjusted for sex and average DPOAE levels from 3-8 kHz.

| Upper bound (90<sup>th</sup> percentile) |  |  |  |  |  |  |  |  |
| --- | --- | --- | --- | --- | --- | --- | --- | --- |
|  | Male |  |  |  | Female |  |  |  |
|  | EFR Stimulus |  |  |  |  |  |  |  |
| Average DPOAE level (dB SPL) | RAM 2 kHz | RAM 4 kHz | RAM 8 kHz | SAM 4 kHz | RAM 2 kHz | RAM 4 kHz | RAM 8 kHz | SAM 4 kHz |
| -15 | 29.39 | 21.91 | 29.33 | 8.16 | 26.69 | 24.34 | 27.37 | 8.99 |
| -10 | 28.85 | 22.97 | 29.34 | 9.39 | 27.03 | 25.21 | 27.74 | 9.70 |
| -5 | 28.30 | 24.03 | 29.35 | 10.63 | 27.37 | 26.09 | 28.12 | 10.42 |
| 0 | 27.76 | 25.09 | 29.35 | 11.86 | 27.71 | 26.96 | 28.49 | 11.14 |
| 5 | 27.21 | 26.15 | 29.36 | 13.10 | 28.05 | 27.83 | 28.86 | 11.86 |
| 10 | 26.67 | 27.21 | 29.36 | 14.33 | 28.39 | 28.70 | 29.23 | 12.58 |
| Lower bound (10<sup>th</sup> percentile) |  |  |  |  |  |  |  |  |
|  | Male |  |  |  | Female |  |  |  |
|  | EFR Stimulus |  |  |  |  |  |  |  |
| Average DPOAE level (dB SPL) | RAM 2 kHz | RAM 4 kHz | RAM 8 kHz | SAM 4 kHz | RAM 2 kHz | RAM 4 kHz | RAM 8 kHz | SAM 4 kHz |
| -15 | 15.11 | 15.51 | 16.53 | 1.24 | 15.35 | 15.75 | 18.18 | 0.70 |
| -10 | 15.94 | 15.68 | 16.63 | 1.57 | 15.95 | 16.24 | 18.79 | 1.14 |
| -5 | 16.77 | 15.84 | 16.73 | 1.90 | 16.55 | 16.73 | 19.39 | 1.58 |
| 0 | 17.60 | 16.01 | 16.83 | 2.23 | 17.16 | 17.22 | 19.99 | 2.02 |
| 5 | 18.43 | 16.17 | 16.93 | 2.56 | 17.76 | 17.70 | 20.60 | 2.46 |
| 10 | 19.26 | 16.33 | 17.04 | 2.89 | 18.36 | 18.19 | 21.20 | 2.90 |

Values are in dB SNR.

### Effect of DPOAE adjustment on EFR normative ranges

In males and females for the RAM 2 kHz, RAM 8 kHz, and SAM 4 kHz stimuli, and females for the RAM 4 kHz stimulus, the lower bound of the normative range decreased as DPOAE average from 3-16 kHz decreased (i.e., as DPOAEs became poorer), although the slope of the lower bound varied across stimuli and between males and females. In contrast, for the male RAM 4 kHz normative range, the lower bound was essentially flat. However, when DPOAE average from 3-8 kHz was used, the lower bounds of all the normative ranges decreased as DPOAE average decreased.

The DPOAE average used for the DPOAE adjustment (3-8 kHz or 3-16 kHz) had only small impacts on most of the normative ranges (**Figure 2** compared to **Figure 3**), although the shape of the normative range changed for the male RAM 4 kHz and female and male SAM 4kHz normative ranges depending on which DPOAE average was used. There were no consistent differences observed across stimuli or sex for the lower bounds calculated using the DPOAE average from 3-16 kHz versus the average from 3-8 kHz. (see **Table 1** versus **Table 2**).

### Effect of sex on EFR normative ranges

There were no consistent differences between the male and female EFR normative ranges (left-hand versus right-hand columns in **Figure 2** and **Figure 3**). Sex-based differences in EFR magnitude were most evident for the RAM EFR 8 kHz normative ranges, where the lower bounds of the normative ranges were elevated by 1.65-4.65 dB in females relative to males, depending on the average DPOAE level. In contrast, the lower bounds of the RAM 2 kHz, RAM 4 kHz, and SAM 4 kHz normative ranges and the upper bounds of the RAM 8 kHz normative ranges were slightly higher in males than females in many cases, depending on the average DPOAE level.

### Effect of carrier frequency and modulation type on EFR normative ranges

Depending on sex and average DPOAE level, the lower bounds of the EFR normative ranges varied from magnitudes of 14.30-19.26 dB SNR for the RAM 2 kHz stimulus, 15.51-18.33 dB SNR for the RAM 4 kHz stimulus, 16.53-21.69 dB SNR for the 8 kHz stimulus, and 0.7-2.90 dB SNR for the SAM 4 kHz stimulus. The lower and upper bounds of the RAM EFR normative ranges were considerably larger than the lower and upper bounds of the SAM EFR normative ranges.

### Report of auditory complaints in the high-risk sample

Details about the auditory complaints reported by the high-risk Veteran sample are shown in **Table 3**. Most Veterans reported more than one auditory complaint.

**Table 3.** Report of auditory complaints in the high-risk sample.

| **Complaint** | **# of males** | **# of females** | **Age in years** | **TFI score** | **SSQ score** |
| --- | --- | --- | --- | --- | --- |
| **Tinnitus only** | 3 | 1 | 42 (5.8) | 20.1 (4.8) | 8.4 (0.6) |
| **Speech perception only** | 5 | 8 | 38.0 (5.6) | ----- | 6.7 (1.3) |
| **DST only** | 1 | 0 | 23 | ----- | 7.6 |
| **Tinnitus + speech perception** | 31 | 12 | 37.9 (6.2) | 42.3 (15.6) | 6.0 (1.4) |
| **Speech perception + DST** | 3 | 1 | 39.5 (4.0) | ----- | 6.1 (1.5) |
| **Tinnitus + speech perception + DST** | 8 | 9 | 38.2 (6.9) | 45.0 (11.8) | 5.8 (1.5) |

The last 3 columns show mean values with the standard deviation in parentheses. DST – decreased sound tolerance; TFI – Tinnitus Functional Index; SSQ – Speech, Spatial, and Qualities of Hearing scale

Tinnitus and speech perception difficulty were the most commonly reported combination. Veterans with multiple auditory complaints tended to report greater tinnitus distress (as indicated by higher TFI scores) and speech perception difficulty (as indicated by SSQ12 score) than Veterans with only one complaint.

### Comparison of high-risk sample to EFR normative ranges

EFR data from individuals in the high-risk sample are plotted relative to the EFR normative ranges in **Figures 2-3**, based on the number of auditory complaints reported (triangles and circles). There was no clear difference in the distribution of EFR magnitudes between Veterans with a single auditory complaint versus Veterans with multiple complaints. The percentage of high-risk participants (both males and females) whose EFR magnitude fell below the lower bound of the normative ranges varied based on the EFR stimulus, with the greatest percentage occurring for the SAM 4 kHz stimulus (36%). The percentage of high-risk participants with EFR magnitudes below the lower bound of the normative range was greater than 10% and the percentage of participants with EFR magnitudes above the upper bound was less than 10% for all EFR stimuli except for RAM 8 kHz when adjusted for average DPOAE level from 3-8 kHz, where 11% of high-risk participants surpassed the upper bound. This suggests that there were different EFR magnitude distributions for the low-and high-risk populations. **Table 4** shows the percentage of high-risk participants who fell above or below the EFR normative ranges.

**Table 4.** Percentage of high-risk sample falling above or below EFR normative ranges adjusted average DPOAE level.

|  | Adjusted for sex and average DPOAE level from 3-16 kHz |  |  |  | Adjusted for sex and average DPOAE level from 3-8 kHz |  |  |  |
| --- | --- | --- | --- | --- | --- | --- | --- | --- |
|  | EFR stimulus |  |  |  |  |  |  |  |
|  | RAM 2 kHz | RAM 4 kHz | RAM 8 kHz | SAM 4 kHz | RAM 2 kHz | RAM 4 kHz | RAM 8 kHz | SAM 4 kHz |
| Below normative range | 21.7% | 22.9% | 12.0% | 35.7% | 26.5% | 22.9% | 12.0% | 35.7% |
| Above normative range | 1.2% | 6.0% | 7.2% | 8.3% | 1.2% | 7.2% | 10.8% | 7.1% |

## Discussion

### DPOAE adjustment may not be necessary for EFRs with a 110 Hz modulation frequency

When considering use of the EFR as a diagnostic indicator of cochlear deafferentation, it is important to consider the potential impacts of OHC dysfunction on EFR magnitude because cochlear deafferentation and OHC dysfunction are expected to co-occur in many cases.

There was no consistent trend across males and females and the different EFR stimuli in terms of the impact of the DPOAE adjustment on the normative ranges. For most conditions (e.g., RAM 2 kHz for both males and females), the lower bounds decreased as average DPOAE level decreased, indicating that smaller EFR magnitudes were associated with smaller DPOAE levels. However, for the male RAM 4 kHz normative range adjusted for average DPOAE level from 3-16 kHz, the lower bound was relatively flat, suggesting that average DPOAE level had no impact on the 10^th^ percentile of the EFR magnitude. Use of the 3-16 kHz DPOAE average versus the 3-8 kHz average resulted in changes in the overall shape of the normative range only for the 4 kHz RAM and SAM stimuli, with the biggest changes in the 90^th^ percentile of the male normative ranges.

Computational modeling suggests that the impact of OHC dysfunction on the RAM EFR differs somewhat based on the carrier frequency, with a 4 kHz carrier considerably less impacted by OHC dysfunction than a 2 kHz carrier (Van Der Biest et al., 2023). This would be expected to result in flatter lower bounds for the RAM EFR 4 kHz normative ranges across average DPOAE level compared to the RAM EFR 2 kHz normative ranges, as is observed for both males and females. However, this does not explain why the upper and lower bounds of the male normative ranges for the RAM and SAM 4 kHz stimuli were considerably more impacted by changing the DPOAE average than the upper and lower bounds of the female normative ranges. One possible explanation for the differing behavior in the male normative ranges is that there was some collinearity between average DPOAE level and EFR magnitude among the males in the low-risk sample. This could occur if the male low-risk participants had greater noise exposure histories than the females, increasing the likelihood of correlated OHC damage and deafferentation. Although the average LENS-Q score was 3.4 for both low-risk males and low-risk females, suggesting similar degrees of lifetime noise exposure, data from mice suggests that males may be more susceptible to noise-induced cochlear damage due to lower levels of estrogen, which protects the cochlea from noise damage (Shuster et al., 2021). If males are more susceptible to noise-induced cochlear damage than females, then cochlear deafferentation and poorer DPOAE levels may be more strongly correlated in the male low-risk participants than the females. This correlation will impact statistical adjustment of the normative ranges for DPOAE levels in the male low-risk participants, making it more challenging to interpret the normative ranges.

This suggests that it is important to carefully consider whether a DPOAE adjustment should be used for the EFR normative ranges. Computational modeling by Encina-Llamas et al. (2019) suggests that subclinical OHC dysfunction in individuals with normal hearing should have limited impact on EFR magnitude. Given that the normative ranges were developed in individuals with normal audiograms, limiting the degree of possible OHC dysfunction, it may not be necessary or advisable to adjust for DPOAE level for these normative ranges. For this reason, EFR normative ranges for males and females that were not adjusted for average DPOAE level were generated (**Figure 4**). These sex-specific normative ranges were calculated using the same quantile regression approach as the DPOAE adjusted normative ranges, but with the DPOAE predictor removed. The values of the upper and lower bounds of these normative ranges are shown in **Table 5** and the percentages of high-risk participants falling outside of the sex-specific normative ranges are listed in **Table 6**.

**Figure 4.**
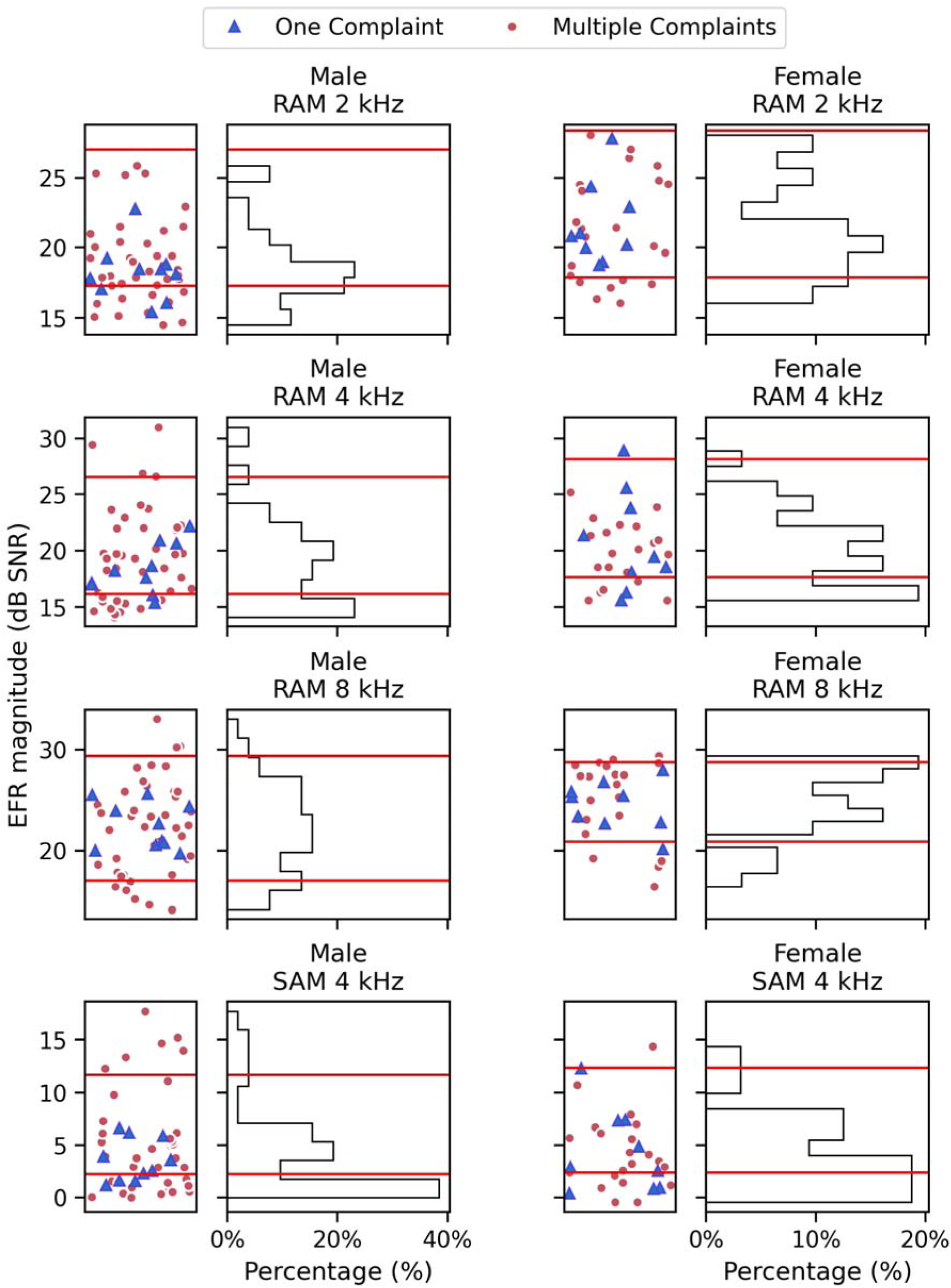
EFR normative ranges adjusted for sex. Each pair of panels shows the normative range for a different EFR stimulus for males on the left-hand side and females on the right-hand side. In the scatterplots, the EFR magnitude for each high-risk participant is represented as a symbol (blue triangle for one auditory complaint, red circle for multiple complaints). Histograms show the distribution of EFR magnitudes for high-risk participants. The upper (90^th^ percentile) and lower (10^th^ percentile) bounds of the normative range are indicated by red horizontal lines on both types of plots.

**Table 5.** Lower and upper bounds for EFR normative ranges unadjusted for DPOAEs.

|  | Males |  |  |  | Females |  |  |  |
| --- | --- | --- | --- | --- | --- | --- | --- | --- |
|  | EFR Stimulus |  |  |  |  |  |  |  |
|  | RAM<br>2 kHz | RAM<br>4 kHz | RAM<br>8 kHz | SAM<br>4 kHz | RAM<br>2 kHz | RAM<br>4 kHz | RAM<br>8 kHz | SAM<br>4 kHz |
| Upper bound<br>(90<sup>th</sup><br>percentile) | 27.03 | 26.59 | 29.35 | 11.67 | 28.40 | 28.16 | 28.75 | 12.35 |
| Lower bound<br>(10<sup>th</sup><br>percentile) | 17.29 | 16.16 | 17.04 | 2.25 | 17.89 | 17.66 | 20.90 | 2.43 |

Values are in dB SNR.

**Table 6.** Percentage of high-risk sample falling above or below EFR normative ranges unadjusted for DPOAEs.

|  | EFR Stimulus |  |  |  |
| --- | --- | --- | --- | --- |
|  | RAM 2<br>kHz | RAM 4<br>kHz | RAM 8<br>kHz | SAM 4<br>kHz |
| **Below**<br>**normative**<br>**range** | 23% | 25% | 14% | 37% |
| **Above**<br>**normative**<br>**range** | 0% | 6% | 6% | 8% |

### No consistent difference in EFR normative ranges between males and females

Although a previous study reported sex differences in the frequency following response (FFR) beginning in adolescence (Krizman et al., 2015), with smaller peak magnitudes for males, no consistent sex differences were observed in the DPOAE-adjusted EFR normative ranges. However, the EFR normative ranges that were not adjusted for DPOAEs showed more consistent sex differences (**Table 5**), with males having smaller magnitudes for the lower bound of the normative range than females for all stimuli and smaller upper bound EFR magnitudes than females for all but the RAM 8 kHz stimulus.

### EFR normative range for a SAM 4 kHz stimulus performed best at differentiating between the low-and high-risk samples

More individuals in the high-risk Veteran sample fell below the sex and DPOAE adjusted EFR normative ranges for the SAM 4 kHz stimulus (36%) than for any other stimuli. This was also true for the EFR normative ranges that were adjusted only for sex, with 37% of high-risk participants falling below the normative range for the SAM 4 kHz stimulus. For the 2 kHz and 4 kHz RAM stimuli, 22-27% of high-risk participants fell below the normative ranges, depending on the DPOAE adjustment that was used. The 8 kHz RAM stimulus performed the most poorly at differentiating between the low-and high-risk samples, with only 12-14% of high-risk participants falling below the normative range.

One potential explanation for the good performance of a 4 kHz carrier is that the 4 kHz cochlear region is particularly vulnerable to noise-induced damage, as indicated by the impacts of noise exposure on the 4 kHz pure tone threshold in Veterans (Wilson & McArdle, 2013). Consistent with this interpretation, Bramhall et al. (2017) found larger mean differences in ABR wave I amplitude between a population at low risk for synaptopathy (young non-Veterans with minimal noise exposure history) and a population at high risk for synaptopathy (young Veterans with a history of military noise exposure) for 4 kHz ABR stimuli than for 1, 3, and 6 kHz stimuli. The good performance of the 4 kHz EFR normative ranges is also consistent with the results of Van Der Biest et al. (2023) where RAM EFR magnitudes for carrier frequencies of 2, 4, and 6 kHz were compared for groups of younger versus older adults. They found significant group differences only for the 4 kHz RAM EFR. Given that Van Der Biest et al. stratified groups by age and excluded participants with high levels of self-reported noise exposure, the ability of the 4 kHz EFR to differentiate groups based on synaptopathy risk factors may not be related to detection of noise-induced cochlear damage.

While it was not surprising to see that a 4 kHz carrier performed well at differentiating between the low-and high-risk populations, it was unexpected to find that the SAM stimulus performed better than the RAM stimulus. Computational modeling (Van Der Biest et al., 2023) suggests that for EFR stimuli modulated at 110 Hz, the RAM stimulus will be more sensitive to synaptopathy than the SAM stimulus. Data from mice with noise-induced and age-related synaptopathy (Buran et al., 2025) indicate that both types of EFR stimuli perform similarly at predicting synapse numbers (for f_m_=110 Hz).

One possible explanation for the better performance of the SAM stimulus is that substantially more trials were collected for the SAM EFR (3002) than for the RAM EFR (1002) because the SAM EFR has a smaller magnitude than the RAM EFR and requires more averaging to get a robust response. The additional averaging lowers the noise floor by 20 • log_10_(√3), or 4.7 dB, which may make the SAM EFR more sensitive.

Another possible explanation is the difference in calibration of the RAM and SAM stimuli, which resulted in an RMS level that was-6 dB lower for the RAM stimuli than for the SAM stimuli. Computational modeling suggests that the contribution of low and medium spontaneous rate fibers to EFR magnitude is greater for a 70 dB SPL stimulus than for a 64 dB SPL stimulus (Verhulst et al., 2018). Given that low spontaneous rate auditory nerve fibers appear to be preferentially impacted by age (Schmiedt et al., 1996) and noise exposure (Furman et al., 2013), this stimulus level difference could give the SAM normative ranges an advantage in terms of differentiating between the low-and high-risk populations. However, computational modeling suggests that high spontaneous rate fibers dominate the EFR response (Encina-Llamas et al., 2019) and would be relatively unimpacted by this change in stimulus level (Verhulst et al., 2018).

### Clinical Implications

This study provides clinicians with normative ranges that can be used to interpret EFR magnitudes measured in response to several different stimuli. If a patient with a normal audiogram has a measured EFR magnitude that falls below the EFR normative range, this suggests that the patient has a significant degree of cochlear deafferentation. This finding will guide targeted interventions for monitoring and treatment, including counseling, hearing conservation programs, amplification strategies, and future pharmaceutical treatment.

Of the stimuli that were evaluated in this study, the SAM EFR for a 4 kHz carrier appears to be the best stimulus to use for this purpose because the SAM 4 kHz normative ranges identified the highest percentage of individuals from the sample at high-risk for synaptopathy as having abnormally small EFR magnitudes. However, the test time for collecting the SAM EFR is 20 minutes longer than for the RAM EFR, which may limit clinical feasibility. Although RAM EFR normative ranges for 2 kHz and 4 kHz did not perform as well as the 4 kHz SAM EFR normative ranges, they identified 22-27% of the high-risk participants as having abnormally small EFR magnitudes. These normative ranges could be used in situations where time does not permit measurement of the SAM EFR.

Adjustment for DPOAEs did not impact the lower bounds of the normative ranges in a consistent manner and computational modeling suggests that this adjustment may not be needed in individuals with normal audiograms. Given these results, it is recommended that clinicians use the EFR normative ranges that are not adjusted for DPOAEs (**Figure 4** and **Table 5**) to identify patients with significant degrees of cochlear deafferentation. However, it is important to note that the normative ranges were developed in individuals with normal audiograms. For this reason, the EFR normative ranges presented in this report should only be used to identify cochlear deafferentation in patients with normal audiograms, where OHC dysfunction should be minimal.

Given that measurement systems may vary, particularly across manufacturers, it is recommended that clinicians validate the EFR normative ranges in a low-risk sample tested with their own equipment prior to using them for diagnostic purposes. Sample characteristics should mirror those for the low-risk sample used in this study in terms of age, reported noise exposure, and lack of auditory complaints. In a new low-risk sample, approximately 10% of individuals should have EFR magnitudes above the normative range and 10% should have EFR magnitudes that fall below the normative range. To facilitate this process, the **Supplemental Data** include all.wav files and IHS.STM files for the EFR stimuli used in the study.

### Limitations

A recent study suggests that it may be common for post-auricular muscle (PAM) artifact to confound FFR measurements, particularly when the reference electrode is placed on the mastoid, resulting in artificially large FFR magnitudes (Bidelman et al., 2024). The PAM is a sound-evoked muscle response that is enhanced by tension and flexion of the neck muscles (Hall, 1992). Bidelman et al. found that PAM artifact was greatest when eyes were gazing to the left or right and for lower frequency stimuli (e.g., 100 Hz). Given that the EFR is recorded in a similar manner to the FFR, there is also potential for PAM contamination of EFR measurements, leading to inflated EFR magnitudes. In this study, the EFR was measured while participants lay flat in a recliner with their head and neck well supported and closed their eyes, limiting neck tension and flexion and prolonged shifts in eye gaze. In addition, use of 2-8 kHz EFR carrier frequencies should reduce the impact of PAM on the EFR measurements. To evaluate whether PAM artifact may have impacted the study results, raw EFR recordings from each participant were evaluated for peaks occurring 12.5-15 msec after the stimulus onset (the expected PAM latency; O’Beirne & Patuzzi, 1999). See the **Supplemental Data** for a detailed description of the PAM analysis. Eight participants (six from the low-risk normative sample and two from the high-risk comparison sample) were identified as having suspected PAM in their EFR. A reanalysis of the sex-adjusted normative ranges with these participants excluded had little impact on the lower bounds of the normative ranges or the percentage of high-risk participants falling below the normative ranges (no more than 2% change; See **Supplemental Data**). Clinicians who use the EFR normative ranges in the future should ensure that the head and neck are well supported and the eyes are closed when obtaining EFR measurements to limit contamination from PAM. For patients whose EFR measurements exceed the upper bound of the normative range, it is advisable to reposition the patient and then retest the EFR.

Given that the EFR with a modulation frequency of 110 Hz (f_m_ =110 Hz) is expected to be generated primarily by the auditory brainstem (Herdman et al., 2002; Kiren et al., 1994; Kuwada et al., 2002), it is possible that central gain could impact EFR magnitude. Individuals with cochlear deafferentation may develop increased central gain in the auditory brainstem (e.g., Chambers et al., 2016), which could elevate EFR magnitude. In this study, it is possible that contributions from central gain to EFR magnitude resulted in a reduction in the number of individuals from the high-risk sample who fell below the normative ranges. Thus, the EFR normative ranges may identify some individuals as “normal” even when they have considerable cochlear deafferentation. Other physiological measures that are more specific to auditory nerve function, such as ABR wave I amplitude and the EFR (f_m_ =1000 Hz), are less likely to be contaminated by central gain. Data from mice with age-related and noise-induced synaptopathy suggest that ABR wave 1 amplitude and the RAM and SAM EFR (f_m_ =1000 Hz) are better able to predict synapse numbers in cases of broad synaptic loss than RAM and SAM EFR (f_m_ =110 Hz), while ABR wave 1 amplitude performs better than EFR measures (f_m_ =1000 Hz or 110 Hz) for focal synaptic loss (Buran et al., 2025). While it is unclear whether RAM EFR (f_m_ =1000 Hz) can be obtained in humans, use of normative ranges for ABR wave I amplitude or SAM EFR (f_m_ =1000 Hz) may be preferred over EFR (f_m_ =110 Hz) normative ranges to limit the impacts of central gain.

## Conclusions

EFR normative ranges were developed in a population at low risk for cochlear synaptopathy, young adults with normal audiograms and minimal noise exposure history. When individuals from a population at high risk for cochlear deafferentation, Veterans with normal audiograms and auditory complaints, were compared to these normative ranges, the SAM EFR normative ranges for a 4 kHz carrier performed best at differentiating between the low and high-risk samples. Although the EFR normative ranges varied by average DPOAE level, the impacts of the DPOAE adjustment on the lower bounds of the normative ranges were not consistent and computational modeling suggests that this adjustment may not be needed in individuals with normal audiograms. Sex-specific 4 kHZ SAM EFR normative ranges, unadjusted for DPOAE level, can be used by clinicians and researchers in the future to identify individuals likely to have high degrees of cochlear deafferentation.

## Data Availability

The datafiles generated during and/or analyzed during the current study are available from the corresponding author on reasonable request.

## Acknowledgements

This work was supported by the Department of Veteran’s Affairs, Veterans Health Administration, Rehabilitation Research and Development Service – Award #C3804R/I01 RX003804 (to N.F.B) and by resources and facilities at the VA National Center for Rehabilitative Auditory Research (NCRAR) [Center Award #C2361C/I50 RX002361] at the VA Portland Health Care System in Portland, OR. The opinions and assertions presented are private views of the authors and are not to be construed as official or as necessarily reflecting the views of the Department of Veterans Affairs.

