## Supplemental Data for "Use of Envelope Following Response Normative Ranges for Diagnosing Cochlear Deafferentation"

**Detailed description of analysis used to identify PAM in the EFR recordings**

First, the raw 4 kHz RAM EFR magnitudes (i.e., not referenced to the noise floor) were evaluated for each participant. One participant had an EFR magnitude that was twice as large as the next-largest amplitude observed. For that participant, there were peaks roughly 15 msec after the stimulus onset in the average EFR waveform. These peaks were very uncommon for other participants, with almost none showing a clear, consistent peak within the expected time window for PAM (12.5 to 15 msec). Based on these observations, a quantitative criterion was established for identifying potential PAM artifacts in the average EFR waveform. The average peak amplitude within the 12.5 to 15 msec window relative was calculated across all 55 cycles in the 500 msec stimulus and called putative PAM (P_PAM_). P_PAM_ was compared to a reference distribution (Ref) of 55 peak amplitudes from randomly-selected timepoints throughout the waveform. The fraction of the Ref distribution that was exceeded by P_PAM_ was calculated to establish whether P_PAM_ was much larger in magnitude than points randomly chosen throughout the waveform (F_Ref_). The probability that a given participant’s P_PAM_ was an outlier was calculated under the assumption that F_Ref_ and P_PAM_ follow a joint normal distribution. Participants where the probability density function of the joint distribution of F_Ref_ and P_PAM_ was less than 0.05 were identified as having suspected PAM.

|  | **Males** | | | | **Females** | | | |
| --- | --- | --- | --- | --- | --- | --- | --- | --- |
|  | **EFR Stimulus** | | | | | | | |
|  | **RAM 2 kHz** | **RAM 4 kHz** | **RAM 8 kHz** | **SAM 4 kHz** | **RAM 2 kHz** | **RAM 4 kHz** | **RAM 8 kHz** | **SAM**  **4 kHz** |
| **Upper bound (90^th^ percentile)** | 27.31 | 26.59 | 29.36 | 12.8 | 28.45 | 27.50 | 28.75 | 12.35 |
| **Lower bound (10^th^ percentile)** | 16.99 | 16.16 | 16.77 | 1.94 | 17.57 | 17.66 | 20.82 | 2.39 |

**Supplemental Data Table 1. Lower and upper bounds for EFR normative ranges unadjusted for DPOAEs after removing participants with suspected PAM.**

Values are in dB SNR.

**Supplemental Data Table 2. Percentage of high-risk sample falling above or below EFR normative ranges unadjusted for DPOAEs after excluding participants with suspected PAM.**

|  | **EFR Stimulus** | | | |
| --- | --- | --- | --- | --- |
|  | **RAM 2 kHz** | **RAM 4 kHz** | **RAM 8 kHz** | **SAM 4 kHz** |
| **Below normative range** | 21% | 26% | 14% | 37% |
| **Above normative range** | 0% | 4% | 5% | 6% |
